# Functional genomic dissection of MS risk loci reveals convergence of cis and trans gene regulatory mechanisms in microglia

**DOI:** 10.1101/2025.06.16.25329681

**Authors:** Michael D. Gallagher, Xochitl Luna, Wenjuan Du, Kathryn E.A. Hazel, Moritz List, Zeynep Aydin, Orian Stapleton, Yiran Cheng, Bingbing Yuan, Keya Viswanathan, Heather R. Keys, George W. Bell, Dheeraj Malhotra, Richard A. Young, Rudolf Jaenisch, Olivia Corradin

## Abstract

Neurological diseases (NDs) are a major source of unmet medical need, and translational insights have been hampered by complex underlying pathophysiologies and limitations of experimental models. Noncoding single nucleotide polymorphisms (SNPs) at hundreds of loci have been linked to ND risk by genome-wide association studies (GWAS), but the causal genes and pathways are largely unknown. Despite the multicellular pathology of complex traits like multiple sclerosis (MS), functional studies that aim to characterize the molecular impact of disease-associated SNPs often investigate all SNPs linked to disease in the same cellular context. Here, we combine a computational approach to predict the pathogenic cell type of individual risk loci with functional CRISPR perturbation studies in iPSC-derived microglia cells (iMGLs). ND SNP enrichment in cell type-specific enhancers is similar between primary and iPSC-derived cells, and mechanistically supported by shared enhancer-promoter interactions. We apply a novel Perturb-seq platform to interrogate MS risk SNPs in iMGL, identifying likely *cis*-acting causal risk genes at 5 of 9 loci, as well as downstream differentially expressed genes (DEGs). Despite being found in *trans* to MS risk SNPs, downstream DEGs are substantially enriched for MS heritability. Downstream DEGs from all 5 target genes show significant overlap, converging on genes related to cytokinesis, phagocytosis, and mitochondrial metabolism. We then compared downstream DEGs to gene expression patterns observed in MS patient tissue studies and observed marked similarities, demonstrating that genes dysregulated as the result of GWAS loci perturbation mirrored effects observed in microglia found in MS patient lesions. Collectively, these results demonstrate that cell type aware functional studies can be used to translate ND SNP associations into mechanistic insights and reveal novel convergent biological mechanisms underlying complex traits.

## Main Text

The pathogenesis of complex traits such as multiple sclerosis (MS) frequently involves multiple cell types and organ systems. MS is both an autoimmune and neurodegenerative disease, characterized by immune cell infiltration into the central nervous system (CNS) leading to a loss of myelin and neurons as the disease progresses [1]. Immune cells such as B and T lymphocytes, as well as glial cells, including oligodendrocytes and microglia, have been implicated in disease pathogenesis. GWA studies have identified more than 200 risk alleles associated with MS, the majority of which are found in the noncoding genome [2]. While these loci are globally enriched for B and T cell active enhancers, the cell type affected by each individual risk allele remains largely unknown [3,4]. We previously devised an approach, termed ‘outside variant analysis’, to identify the cell type that is critically affected by the presence of a given disease-associated allele [5,6]. Here, we extend the approach to identify noncoding MS risk loci that contribute to disease through gene dysregulation in microglia and leverage a microglia-optimized Perturb-seq [7],[8] platform to identify both the cis and trans effects of enhancers associated with MS risk on gene expression.

### Outside variant analysis identifies 17 MS risk loci predicted to influence microglia function

The outside variant approach leverages analysis of genetic risk information of SNPs that physically interact with the same target promoter and compares intralocus genetic risk patterns to chromatin activity to predict ‘pathogenic cell types’ i.e. the cell type in which the risk allele functionally contributes to disease pathogenesis (Fig. 1A). We utilized Promoter Capture Hi-C (PCHi-C) data from 15 immune cell types [9] as well as H3K4me3 PLAC-seq interaction data from primary microglia, astrocytes, oligodendrocytes and neurons [10] to identify putative gene targets of 200 published MS GWAS loci. We identified candidate ‘outside variants’, genetic variants that are physically linked to the same target promoters as MS GWAS loci and identified their impact on genetic risk as previously described (see methods). We then compared local genetic risk patterns and H3K27ac activity in each cell type to predict pathogenic cell types. Cell types in which outside variants that alter MS risk have a high concordance with H3K27ac enrichment patterns are predicted as likely pathogenic (Fig. 1B). We predicted pathogenic cell types for 141/200 MS risk loci (Fig. 1C). A minority of loci (n=18) had similar H3K27ac activity across immune and neural cell types, and an additional 9 had similar activity across immune cells. For the remaining 114 MS risk loci, cell type or lineage-restricted predictions were identified. Notably, this included 17 MS risk loci predicted to act pathogenically in microglia (Fig. 1D). Amongst these predictions is rs57116599, linked to the gene *MERTK*. *MERTK* has previously been shown to be required for microglia activation and plays a key role in clearing myelin debris to enable remyelination [11]. To further assess the validity of these predictions, we performed stratified linkage disequilibrium score regression (s-LDSC) [12] to compare cell type predictions to the heritability of 55 complex traits. MS risk loci predicted to act in immune cells showed greater heritability enrichment for other autoimmune traits such as primary biliary cirrhosis, compared to MS risk loci predicted to act in CNS cell types (Fig 1E). MS risk loci predicted to act in neurons had the strongest enrichment of generalized anxiety disorder, and loci predicted to act in microglia had the strongest enrichment for Alzheimer’s disease. We previously demonstrated that loci predicted to act in oligodendrocytes inhibited oligodendrocyte maturation and the generation of new myelin [5]. Collectively, these results support the outside variant approach predictions and the pursuit of functional studies in the predicted cell type.

**Figure 1:**
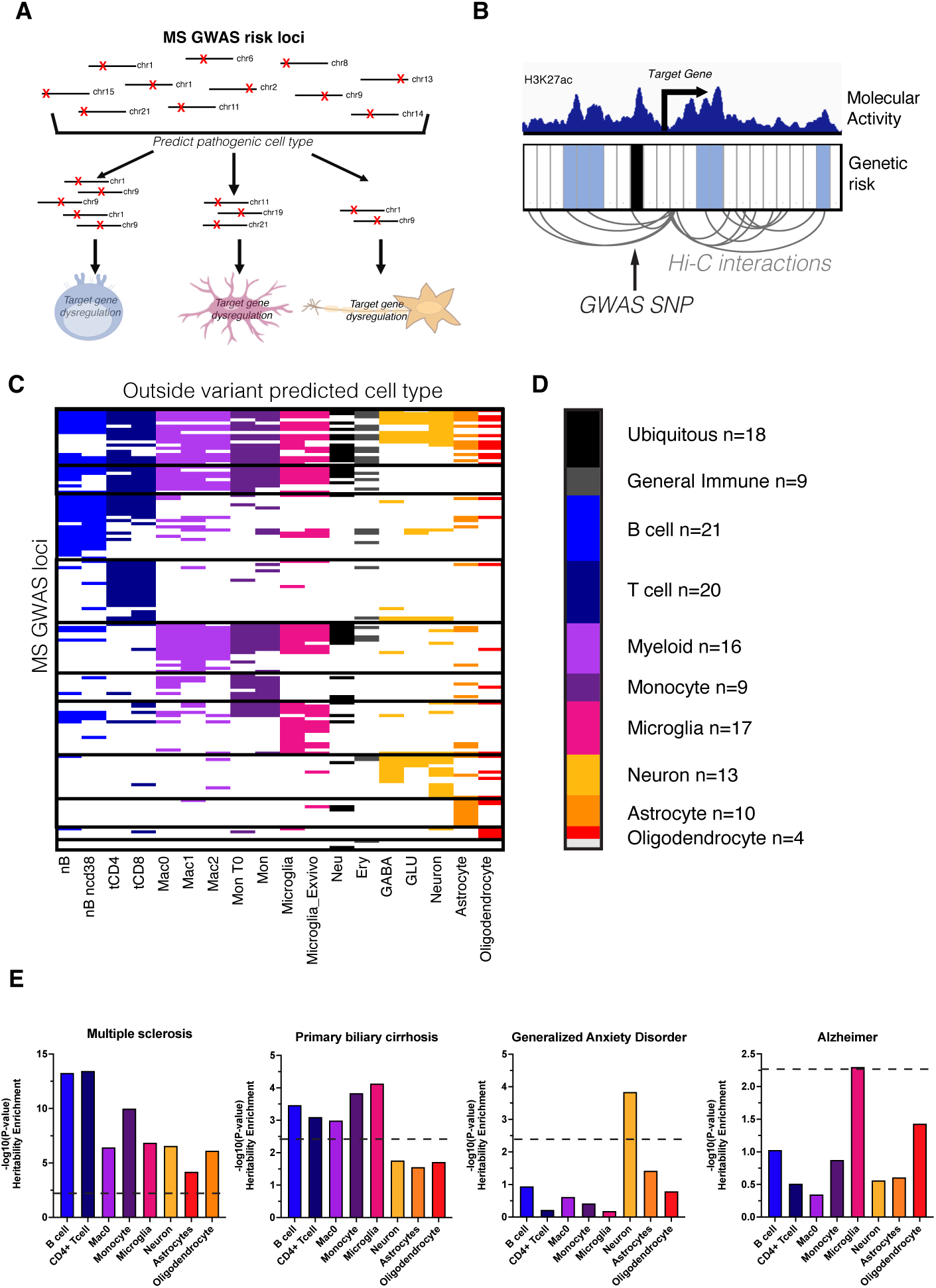
Outside variant analysis identifies 17 MS risk loci predicted to influence microglia function. **(A)** Schematic overview. Individual GWAS loci are segregated to the cell type in which they exert a risk-conferring effect through computational comparison of genetic risk and chromatin activity. **(B)** Diagram showing outside variant method. (Below) Barcode represents genomic regions with outside variants. Regions shaded in blue contain outside variants that significantly alter clinical risk. Variants in unshaded regions do not alter clinical risk. (Top) Example H3K27ac track showing concordance with risk-conferring variants and chromatin activity. Cell types with strong concordance between H3K27ac and outside variants that alter clinical risk are predicted as a likely pathogenic cell type. **(C)** Heatmap showing outside variant cell type predictions generated for MS GWAS loci (N=141/200). Each row represents a GWAS locus and the column is shaded if the cell type listed on the x-axis is predicted. **(D)** Number of loci predicted per cell type or cell group in (C). **(E)** S-LDSC heritability enrichment across MS GWAS loci separated by predicted cell types. Dashed line shows P-value threshold after multi-test correction for 10 evaluated cell type groups shown in (D).

### Human iPSC-derived microglia recapitulate ex vivo microglia at disease-associated loci

We next sought to evaluate whether iPSC-derived CNS cell types could be leveraged to investigate the functional consequence of predicted MS risk loci. We generated neural progenitor cells (NPCs), excitatory neurons (iPSC-N), astrocytes (iPSC-A) and microglia (iMGL) from female and male iPSCs, using published and modified protocols [13–16] (Fig. 2A). Cell identity and function were validated with immunocytochemistry (ICC), transcriptomics and functional assays. NPCs stained positive for PAX6 and Nestin and displayed enrichment of neuronal and ectoderm differentiation gene ontology (GO) pathways (Fig. 2B, Fig. S1C) [17]. iPSC-Ns displayed extensive MAP2+ processes, upregulation of neuronal and synaptic pathways, and spontaneous synaptic activity after 21 days of differentiation (Fig. 2B, S1A-B,D). *NEUROD2* and *NEUROD6* were uniquely expressed in iPSC-Ns, consistent with a cortical glutamatergic identity. iPSC-As displayed typical “star-like” astrocyte morphology, expressed high levels of *S100B*, *GFAP*, *VIM* and *NFIX*, and upregulated the mature astrocyte markers *AQP4* and *ALDH1L1* tens-to hundreds-fold compared to their parental NPCs (Fig. 2B, S1B). GO analyses revealed upregulation of glial cell differentiation and synaptic activity and maturation pathways, consistent with the role of astrocytes in modulating synaptic function [18] (Fig. S1E). iMGL displayed strong GO enrichment of immune-related pathways, including cytokine production, phagocytosis and TYROBP pathways (Fig. S1F), which involve TREM2 (the TYROBP co-receptor)-mediated response to diverse stimuli in brain health and disease [19]. iMGL uniquely expressed the macrophage markers *PU.1*, *IBA1* and *TREM2* (Fig. S1B), and stained positive for IBA1 (Fig. 2B). Finally, these cells demonstrated dose- and actin polymerization-dependent phagocytosis and efficient lipid uptake, consistent with microglial identity and function [20] (Fig. S1H,I).

**Figure 2:**
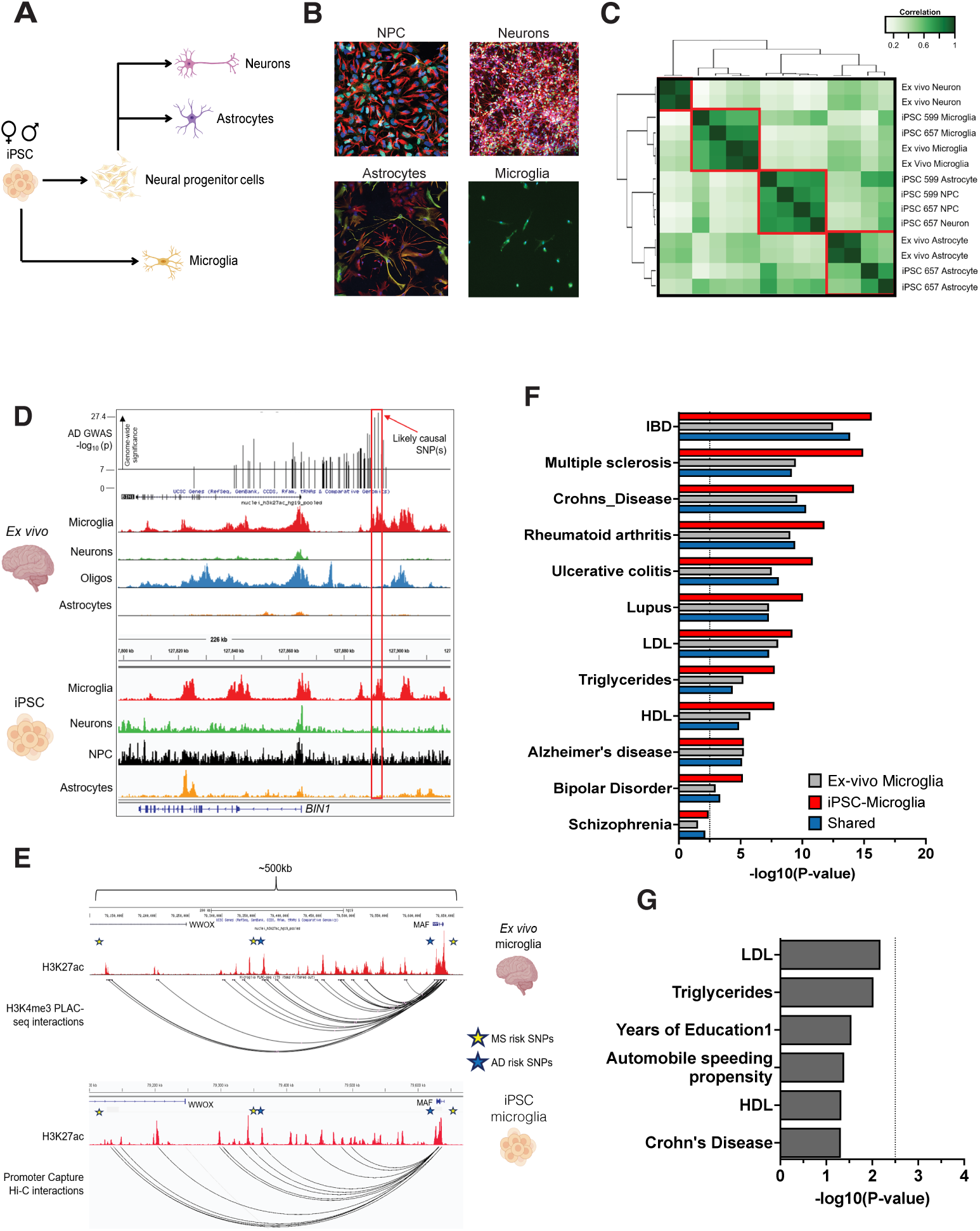
Human iPSC-derived microglia recapitulate epigenetic patterns of ex vivo microglia at disease-associated loci. **(A)** Schematic showing iPSC differentiation into cells of the central nervous system (CNS). **(B)** Fluorescence microscopy images of iPSC-derived cells demonstrate expression of PAX6 (green) and Nestin (red) in NPCs, MAP2 (red), and Ngn2 (green) in neurons, GFAP (green) and Vimentin (red) in astrocytes, and IBA1 (green) in microglia. DAPI is blue. **(C)** Comparison of H3K27ac ChIP-seq profiles from iPSC-derived and ex vivo cells demonstrates clustering of iPSC-derived microglia and astrocytes with their ex vivo counterparts. (**D)** H3K27ac in ex vivo and iPSC-derived cells at the *BIN1* AD risk locus shows overlap of likely causal SNPs (red rectangle) with an experimentally-validated microglia-specific enhancer. AD risk SNPs are indicated by vertical lines above the ChIP-seq tracks, AD GWAS p-value is shown on the y-axis, and the horizontal line indicates the genome-wide significance cutoff. **(E)** PLAC-seq from ex vivo microglia and PCHi-C from iPSC-derived microglia show similar enhancer-gene chromatin contacts near MS and AD risk SNPs. **(F)** S-LDSC SNP heritability enrichments for 12 diseases and traits are shown for H3K27ac-defined enhancers identified in ex-vivo microglia, iMGL, and regions with H2K27ac-enrichment found in both iMGL and ex vivo microglia (shared). **(G)** S-LDSC SNP heritability enrichments for H3K27ac defined enhancers that are identified in ex vivo microglia but are inactive in iMGL.

As ∼90% of GWAS risk SNPs are located in noncoding regions that may alter disease risk through gene regulatory processes [21,22], we performed chromatin immunoprecipitation and high-throughput sequencing (ChIP-seq) for H3K27ac, a marker of active promoters and enhancers [23,24] in all iPSC-derived cells (Fig. S2A). These data demonstrated expected cell type-specific patterns at marker gene loci, including *NEUROD2* in iPSC-Ns, *ALDH1L1* in iPSC-A and *SPI1* (encoding PU.1) in iMGL (Fig. S2B-D). To determine how similar our iPSC-derived cells are to their in vivo counterparts, we compared these data with those from primary, uncultured (ex vivo) neurons, astrocytes, oligodendrocytes and microglia, which were sorted directly from freshly resected human cortical tissue [10]. Notably, these ex vivo data were obtained from primary cells that were never exposed to a cell culture environment, and thus represent a “gold standard” dataset for brain cell type-specific epigenetic patterns. Hierarchical clustering demonstrated the strongest similarity between ex vivo microglia (exMG) and iMGL, moderate similarity between ex vivo astrocytes and iPSC-A, and clustering of NPCs, iPSC-Ns and one iPSC-A replicate away from ex vivo cells (Fig. 2C). The cell type marker loci mentioned above displayed similar H3K27ac patterns in ex vivo cells compared with iPSC-derived cells (Fig. S2).

After observing the strongest correlation between iMGL and exMG we sought to investigate the extent to which iMGL mirror exMG at disease-associated SNPs. We performed s-LDSC on H3K27ac-enriched regions identified in iMGL, exMG, as well as regions with H3K27ac enrichment identified in both (shared). We observed similar heritability enrichment patterns between exMG and iMGL enhancers (R=0.88), which are both enriched for SNPs associated with inflammatory bowel disease, multiple sclerosis (MS), Alzheimer’s disease (AD), and several other autoimmune conditions (Fig. 2F). An example of a disease risk locus at which iMGL display similar epigenetic features as exMG is shown in Fig. 2D. The *BIN1* locus is the strongest AD GWAS locus except for APOE [25], with two top risk SNPs overlapping a microglia-specific enhancer. Deletion of this enhancer in iPSCs was reported to reduce BIN1 expression in iPSC-derived microglia, but not neurons or astrocytes [10], suggesting that one or both SNPs may influence AD risk by altering BIN1 levels specifically in microglia.

iPSC-derived cells are thought to resemble fetal rather than adult cells, and be less functionally mature than their *in vivo* counterparts [26]. In addition, microglia and other macrophages are very sensitive to their environments, and rapidly lose tissue-specific *in vivo* enhancers and gene signatures in cell culture environments [27]. To further scrutinize the physiological relevance of our iMGL, we first examined the relationship between context-specific enhancers and disease risk SNPs. Enhancers identified in exMG that are not active in iMGL did not display SNP enrichment for any traits, suggesting where exMG and iMGL differ is not substantially linked to disease biology. [27,28], (Fig. 2G). By assigning these enhancers to their putative target genes using H3K4me3 PLAC-seq data from the same dataset, we observed that the largest loss of H3K27ac signal occurred at the *SALL1* locus, consistent with other studies implicating *SALL1* as an important driver of the homeostatic microglial gene program in vivo [29]. Ex vivo-specific enhancers interacted with genes enriched in Fc-epsilon receptor signaling, regulation of glycogen biosynthesis, cell junction disassembly, and other pathways (Fig. S3C). Thus, while iMGL exhibit epigenetic differences with exMG that are consistent with previous reports, iMGL appear to recapitulate enhancer activity patterns observed in tissue-derived microglia at ND risk loci. Next, we compared the transcriptomic profiles of our iMGL with those from four published protocols, as well as two independent exMG datasets [29–34]. Our iMGLs clustered as or more closely with exMG than iMGLs derived by other differentiation protocols, including the protocol we modified for this study[29]. (Fig. S3A). These results are likely not specific to the exMG dataset used, as two independent exMG datasets cluster very closely together (Fig. S3,B). One explanation for the exMG-like gene signature of our iMGL may be the use of our previously published microglia maturation media, which is fine-tuned to match solute concentrations observed in human cerebrospinal fluid [31].

Finally, as the presence of shared enhancers between exMG and iMGL does not necessarily imply that these enhancers also share target genes, we performed PCHi-C on iMGL to assign putative target genes to microglial enhancers genome-wide. As an example, we highlight the locus *WWOX/MAF* (Fig. 2E) which displays multiple independent sets of SNPs that influence risk for MS and Alzheimer’s disease separately, suggesting pleiotropic mechanisms at this locus. While previous studies have suggested that *WWOX* may be the causal gene at this locus [35], these SNPs, as well as MS risk-modifying outside variants, overlap an extensive network of enhancers spanning ∼500kb that display strong PCHi-C and PLAC-seq interactions with MAF. The epigenetic landscape at this locus is strikingly similar between exMGs and iMGLs, consistent with physiological risk SNP mechanisms in our iMGL model.

Overall, we identified ∼190,000 promoter-based interactions, and found that 96% of all intra-chromosomal (*cis*) interactions spanned less than 1Mb, consistent with predicted enhancer-promoter (E-P) interaction distances (Fig. S3D-E) [36]. There was a median of 3 (mean=4.3) cis interactions per promoter (Fig. S3D), with ∼53% representing promoter-other (P-O) interactions, which likely consist of interactions with enhancers, CTCF sites, and other genomic regions, and ∼47% representing promoter-promoter (P-P) interactions. Promoter-based interactions were significantly and positively correlated with the mRNA levels of the promoter-associated gene (P=1.20×10-137) (Fig. S3F). Furthermore, the distal interacting regions were enriched ∼14X for H3K27ac (Fisher’s exact test, P≈0), suggesting that many are E-P interactions. Finally, the 300 genes with the most interactions were more strongly associated with microglial gene sets than those of any other cell type (Fig. S3G). Overall, these results suggest that our iMGL are physiologically relevant microglial cell models, and well-suited for studying disease-associated gene regulatory mechanisms.

*Adaptation of CRISPRi single cell screen to MS risk loci in iMGL* In order to investigate the mechanisms at MS GWAS loci, we developed a novel CRISPRi single cell screening (Perturb-seq [7,8]) approach, which allows for the identification of SNP enhancer target genes as well as transcriptome-wide effects without inducing double-strand breaks.

However, to determine the effects of CRISPRi on iMGL without the confounding effects of dCas9 activity during iPSC differentiation, sgRNAs and dCas9 cannot both be active before differentiation is complete. While sgRNA libraries can be delivered to iPSCs by lentivirus, this approach requires tight control of inducible dCas9 transgenes, and library representation can become skewed during prolonged differentiations [20]. Therefore, we developed a strategy to perform CRISPR single cell screens in iMGL by combining Vpx-associated [37,38] lentiviral sgRNA delivery with constitutive dCas9 expression. We generated iPSCs containing a dCas9-KRAB transgene in the *AAVS1* safe harbor locus, using the ZIM3 KRAB domain, due to its superior CRISPRi activity [39,40]. After identifying clones with successful integrations, we selected a karyotypically normal clone with robust dCas9-KRAB expression (Fig. S4A-D). After differentiation to iMGL, we tested transduction with a Cas9/EGFP-expressing lentivirus with or without co-packaged Vpx protein. 10 days post-transduction, EGFP was almost completely absent in iMGLs transduced without Vpx, but was visible in most cells transduced with Vpx-associated lentivirus (Fig. S4E). Transduction efficiencies of up to ∼90% were not associated with any cell death, and while modest increases of a subset of secreted pro-inflammatory cytokines were initially observed, they largely recovered over the next several days (Fig. S4F). Similar results were obtained with a published Perturb-seq vector [41] (Fig. 3D, S4G), suggesting that Vpx-based lentiviral transduction is an effective and versatile strategy for delivering gene editing reagents to iMGLs. Next, we assessed CRISPRi activity in our dCas9-KRAB iMGLs by transducing them with Perturb-seq lentivirus containing guides targeting the transcriptional start sites (TSS) of four genes spanning a ∼60-fold range in expression levels in iMGL. These guides achieved ∼50-90+% reduction in mRNA levels by D7 post-transduction (Fig. 3B). To validate CRISPRi activity at enhancers, we targeted the previously studied *BIN1* AD risk SNP enhancer with two guides, and observed ∼20-25% reduction of *BIN1* levels by D7 (Fig. 3B). As enhancer CRISPRi is expected to provide weaker effects than TSS CRISPRi [42], and these magnitudes compared favorably to published iMGL CRISPRi experiments, we used this system for the Perturb-seq screen [10,42].

**Figure 3:**
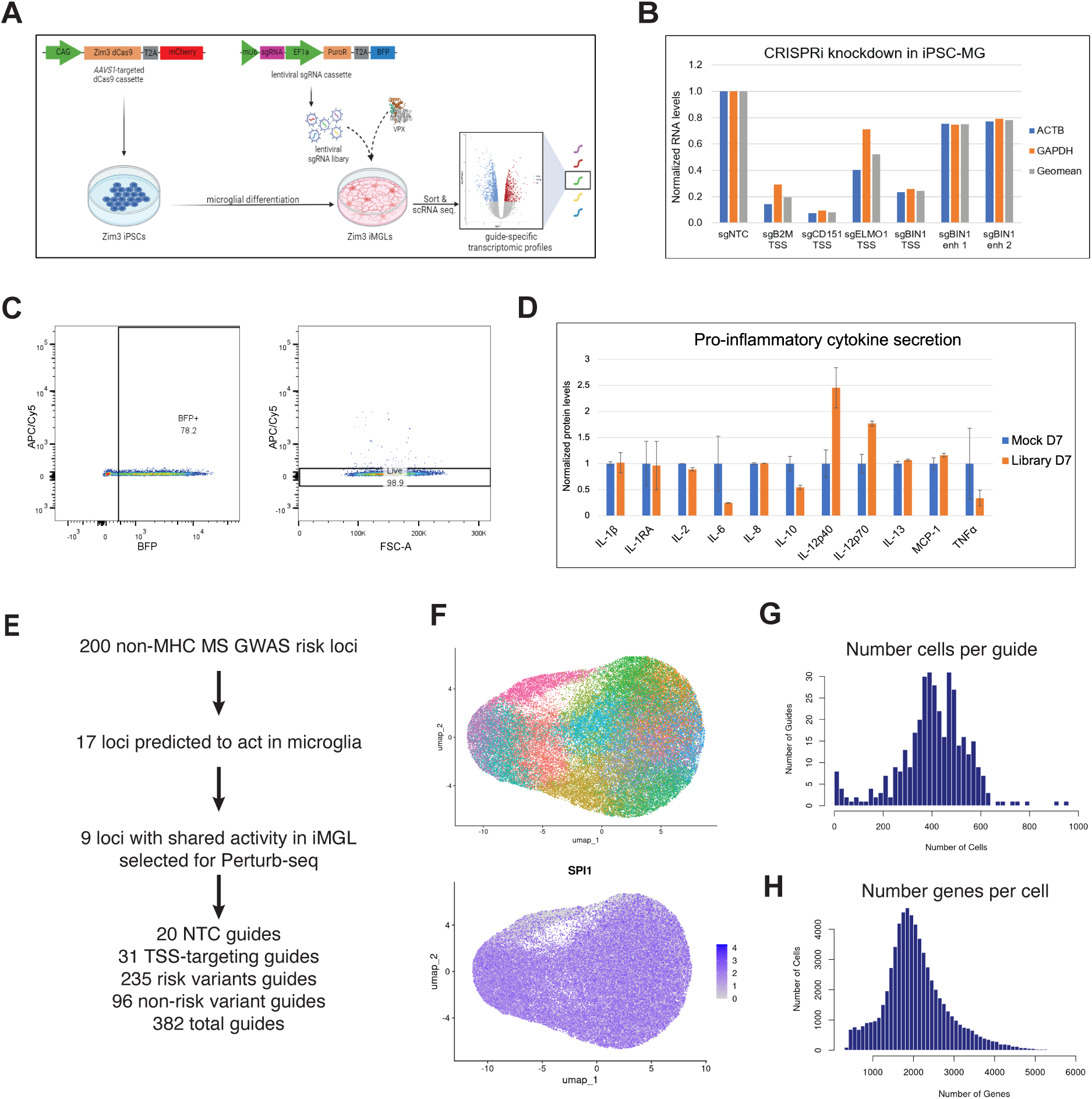
Microglia-compatible Perturb-seq. **(A)** Perturb-Seq experimental design. dCas9-KRAB (ZIM3) iPSCs are differentiated into iMGL, transduced with a lentiviral sgRNA library co-packaged with Vpx, and sorted for scRNA-seq of transduced cells. **(B)** CRISPRi knockdown of genes using TSS-or enhancer-targeting sgRNAs, determined by qPCR. mRNA levels were normalized to. *ACTB*, *GAPDH* or both (Geomean) are shown. **(C)** iMGLs transduced with the Perturb-seq sgRNA library 7 days prior were 78.2% BFP+ and 98.9% viable immediately before scRNA-seq. **(D)** Pro-inflammatory cytokine ELISA analysis of iMGLs transduced with Perturb-seq sgRNA library, compared with untransduced cells otherwise treated identically. **(E)** Flowchart demonstrating selection of loci for Perturb-Seq and sgRNAs cloned into library. **(F)** UMAP (principal components 1-5) shows homogenous microglial population in Perturb-Seq screen (top). Feature plot of *SPI1* (PU.1) expression, a master microglial transcription factor (bottom). **(G)** Histogram of the number of cells expressing each sgRNA. **(H)** Histogram of the number of genes detected per cell.

To select loci for our screen, we compared the 17 MS risk loci we predict to act in microglia to iMGL. We prioritized loci that displayed H3K27ac enrichment at MS GWAS SNPs and risk-conferring outside variants in both exMG and iMGL, but not in other brain cell types. Then we prioritized loci in which the ChIP-seq data showed stronger H3K27ac in microglia than other myeloid enhancers. From this analysis, we selected 9 MS risk loci that display epigenetic evidence of risk SNP mechanisms in microglia, as well as *BIN1* as a positive control for microglia E-P interactions. We designed a sgRNA library containing 3-4 sgRNAs targeting the TSS of the top candidate target gene at each locus, 5 sgRNAs targeting each enhancer overlapping either an MS risk SNP or outside variant SNP, 17 non-targeting controls, and negative controls targeting regions that serve as controls for distance to the nearest gene, location in gene introns, overlap with H3K27ac is non-microglial cell types, or overlap with SNPs not associated with MS risk (Fig. 3E). Overall, our library contains 387 sgRNAs, 382 of which were successfully cloned into our Perturb-seq vector.

We transduced iMGL with this library at an MOI of ∼1.5, and confirmed that transduction was non-toxic and well tolerated (Fig. 3C-D). We prepared seven 10X 5’GEM-X scRNA-seq libraries with the CRISPR feature barcode, which provides an sgRNA-specific RT-PCR primer such that sgRNA expression can be directly sequenced in every cell [41]. We generated ∼10 billion read pairs and single cell transcriptomes for ∼120,000 cells with assigned sgRNAs. scRNA-seq revealed high homogeneity in the transcriptomes of transduced iMGL, including homogeneous expression of the microglia markers *SPI1* (PU.1), *AIF1* (IBA1), and *TREM2*, with more variable expression of the homeostatic marker *TMEM119*. The specificity of our iMGL cell identity was supported by undetectable *CD34*, a hematopoeitic progenitor marker, and near zero levels of *LYVE1*, a marker of brain border-associated macrophages that are otherwise highly similar to microglia [43] (Fig. 3F, Fig S5). We were able to assign sgRNAs to 66% of sequenced cells. After filtering for high-quality transcriptomes, a median of 414 cells for each sgRNA were recovered, with a median of 1,965 detectable genes per cell (Fig. 3G-H, methods).

### Identification of cis target genes for 5/9 microglia risk loci

While chromatin interaction data can reveal putative target genes of enhancer loci, there are frequently multiple enhancers and multiple genes implicated by identified interactions. Thus, we first leverage our Perturb-seq dataset to investigate the cis target genes of each GWAS locus. We performed individual sgRNA analysis, in which differentially expressed genes were identified by comparing cells containing each individual guide to cells receiving only non-targeting control (NTC) guides (N=1,021). We then evaluated the impact of each sgRNA on all genes within 1Mb of the sgRNA to identify cis target genes. As an example, we highlight the *MERTK* locus (Fig 4A). SNPs in strong linkage disequilibrium with GWAS SNPs in this region extend across a large genomic regions (134 kb) encompassing the gene body of *MERTK*. We identified 3 enhancer regions with GWAS SNPs or risk-conferring outside variants in this locus, largely found in intronic regions of *MERTK*. sgRNA targeting each of these enhancers led to modest (43%) but significant reduction (Benjamini-Hochberg corrected P=7.8E-13) in *MERTK* expression (Fig. 4B). In comparison, sgRNA targeting the *MERTK* TSS results in a 73% reduction in *MERTK* (corrected P=1.1E-32) (Fig. 4C). Notably, 27/28 sgRNA targeting local control regions did not impact *MERTK* expression.

**Figure 4:**
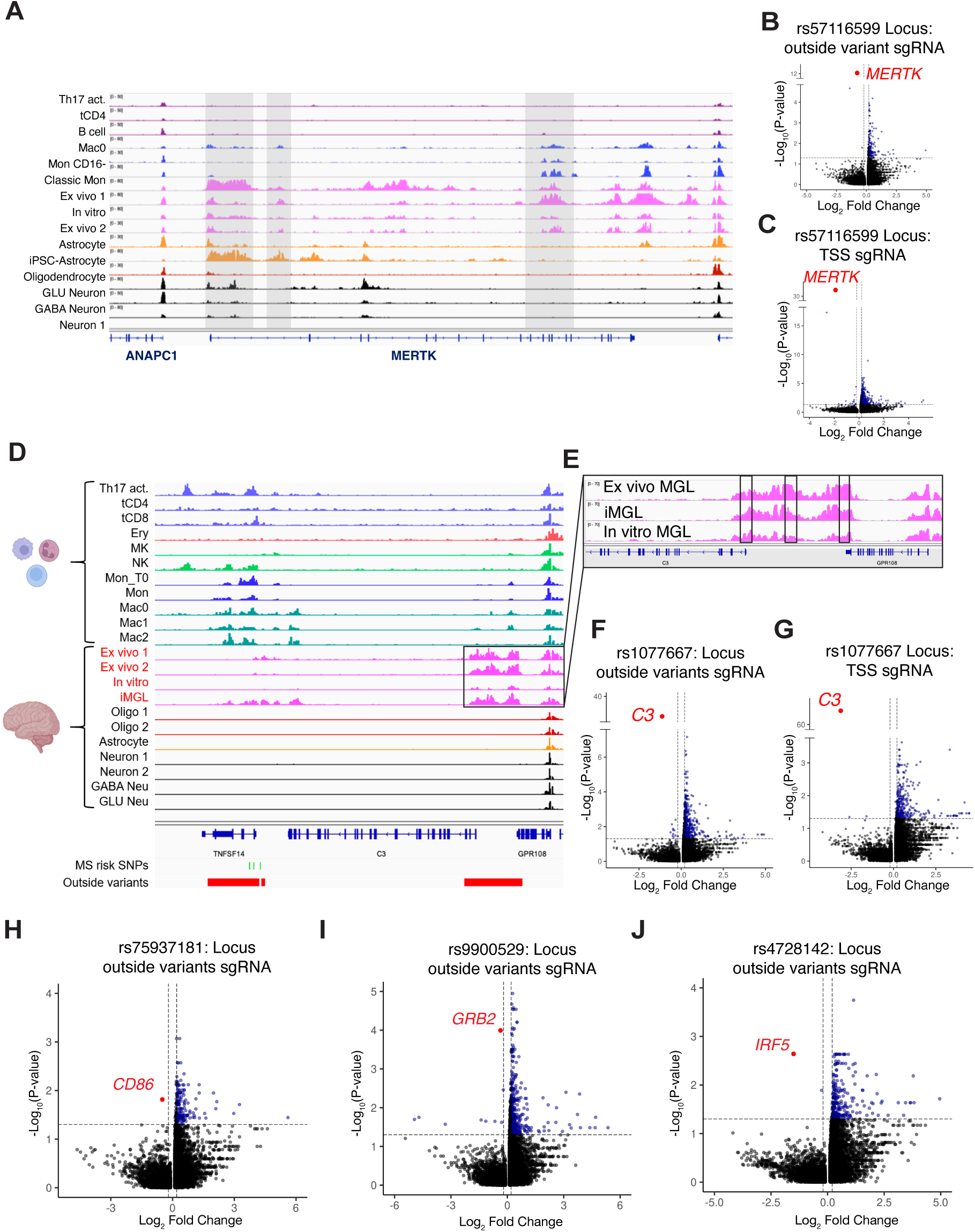
Identification of *cis* target genes for 5/9 microglia risk loci. **(A)** H3K27ac ChIP-Seq data from immune and CNS cells at the *MERTK* locus. Regions shaded in gray contain outside variants that influence MS risk. sgRNAs that target these regions significantly impact *MERTK* expression. **(B)** Volcano plot of differentially expressed genes (DEGs) from *MERTK* distal enhancer CRISPRi. Blue dots exceed 15% fold change and Benjamini-Hochberg-corrected p-value threshold of P<0.05 for all volcano plots. *MERTK* FC=43%, corrected P=7.8E-13. **(C)** Volcano plot of DEGs from *MERTK* TSS CRISPRi. *MERTK* FC=73%, correct P=1.1E-32. **(D)** H3K27ac ChIP-Seq data from immune and CNS cells at the *TNFSF14/C3* locus. Below, in green, MS GWAS SNPs. In red, putative enhancers with outside variants that affect disease risk. **(E)** Zoomed in view of *C3* putative enhancer cluster. Boxes indicate regions with outside variants that influence disease risk and sgRNA target regions that alter *C3* expression. **(F)** Volcano plot of DEGs from *C3* distal enhancer CRISPRi. *C3* FC=55%, corrected P=6.1E-35. **(G)** Volcano plot of DEGs from *C3* TSS CRISPRi. *C3* FC=89%, corrected P=7.7E-68. **(H)** Volcano plot of DEGs from *CD86* distal enhancer knockdown. FC=16% corrected P=0.015. **(I)** Volcano plot of DEGs from *GRB2* distal enhancer knockdown. *GRB2* FC=22%, corrected P=0.00010. **(J)** Volcano plot of DEGs from *IRF5* distal enhancer knockdown. *IRF5* FC=65%, corrected P=0.0023.

We additionally observed multiple functional enhancers at the *TNFSF14*/*C3* locus (Fig 4D). While MS GWAS risk SNPs overlap H3K27ac near the 5’ end of *TNFSF14* in many immune cell types, we identified a set of outside variants that overlap robust microglia-enriched H3K27ac upstream of the gene *C3*, suggesting that risk SNPs at this locus may act through microglia with effects on one or both of these genes. sgRNA targeting three enhancer regions show significant impact on *C3* expression (Fig. 4E), but no impact on proximal genes *TNFSF14* or *GPR108* (Fig 4F). Additionally, sgRNA targeting the reported GWAS SNP region did not have a significant impact on any genes at the locus. This suggests the locus may have diverse effects in different cell types. Alternatively, this may indicate that the causal SNP(s) are distal to the reported MS GWAS SNPs.

We identified at least one distal enhancer that has a significant impact on cis target genes for three additional loci, including rs7593718 (CD86), rs9900529 (SLC25A19/GRB2), rs1077667 (IRF5) (Fig. 4H-J). Two enhancers in close proximity, located ∼50kb upstream of CD86 TSS, led to a significant reduction in CD86; outside variant loci found in the 3’UTR of the gene GRB2, 85kb downstream of the GRB2 TSS were found to significantly reduce GRB2 expression, and gRNAs 300bp upstream of IRF TSS significantly reduced IRF5 expression.

For the remaining four loci, we did not confidently identify a *cis* target gene in iMGL. This included the *ELMO1* locus, which contains multiple clusters of enhancers with MS GWAS and outside variants (Fig S6). Despite robust enhancer activity and strong guide coverage, we did not observe a significant impact on *ELMO1*. The strongest distal enhancer guide had a modest corrected p-value of 0.17. Given the high number of enhancers in this region, this may reflect the need for future combinatorial perturbation strategies to account for multiple enhancer dynamics. For two additional loci, we observed a significant impact on putative *cis* target genes *IFNGR1* and *MARCHF1*. However, the sgRNAs with significant impacts were too proximal to the gene’s promoter to be confidently distinguished from TSS-driven effects; more distal gRNA did not impact expression.

### Downstream DEGs converge on shared biological pathways linked to microglia dysfunction in MS

We next sought to identify the downstream impacts of *cis* target gene dysregulation on microglia transcriptomes genome-wide. In order to maximize power to detect downstream DEGs, we pooled all cells containing a sgRNA that significantly impacted *cis* target gene expression for each of the 5 validated *cis* target genes (pooled-cis). We compared these to NTC as well as a pooled control population derived from all cells with sgRNAs that did not influence *cis* target gene expression (pooled-control). These comparisons were highly correlated (Fig. S7) (average Pearson correlation R=0.875, P<2E-7). Pooled-cis vs. pooled-control had the largest number of significant DEGs, suggesting power was the primary distinguisher.

We identified a total of 935 downstream DEGs across all 5 loci. To assess whether these downstream transcriptional changes were relevant to the pathology of MS, we performed s-LDSC. We observed a significant enrichment for MS heritability in downstream DEGs. This enrichment was still prominent after excluding the *cis* target gene loci and the HLA locus (Fig 5A,B). We observed a statistically significant, but more modest enrichment of AD heritability in these DEGs as well, suggesting that these loci may be particularly relevant to MS, rather than merely reflecting general microglia function. We compared downstream DEGs identified across all 5 MS risk loci and identified a striking overlap. 39% of downstream DEGs were identified in two or more *cis* target gene knockdowns and 16% were common to 3 or more, exceeding the overlap expected by chance (permutation P<0.002) (Fig. 5C). The consistency in the downstream DEGs across loci is highlighted in Fig 5D. Of the 935 downstream DEGs, we identified 168 that were downregulated in all 5 MS *cis* target loci. These were significantly enriched for processes including regulation of GTPase activity and phagocytosis [44–46] (Fig 5E). 260 downstream DEGs were upregulated in all 5 loci and were enriched for cellular respiration and oxidative phosphorylation pathways (Fig 5F). Importantly, when we compared NTC cells to pooled-control cells, we did not observe overlaps with downstream DEGs. Furthermore, we performed random permutation of sgRNA assignments and DEG assessment. In random permutation analysis, we did not observe significant convergence of downstream DEGs. Taken together, these results suggest that the substantial overlap in downstream DEGs and biological pathways at these 5 MS risk loci reflect shared microglia biology linked specifically to MS risk.

**Figure 5:**
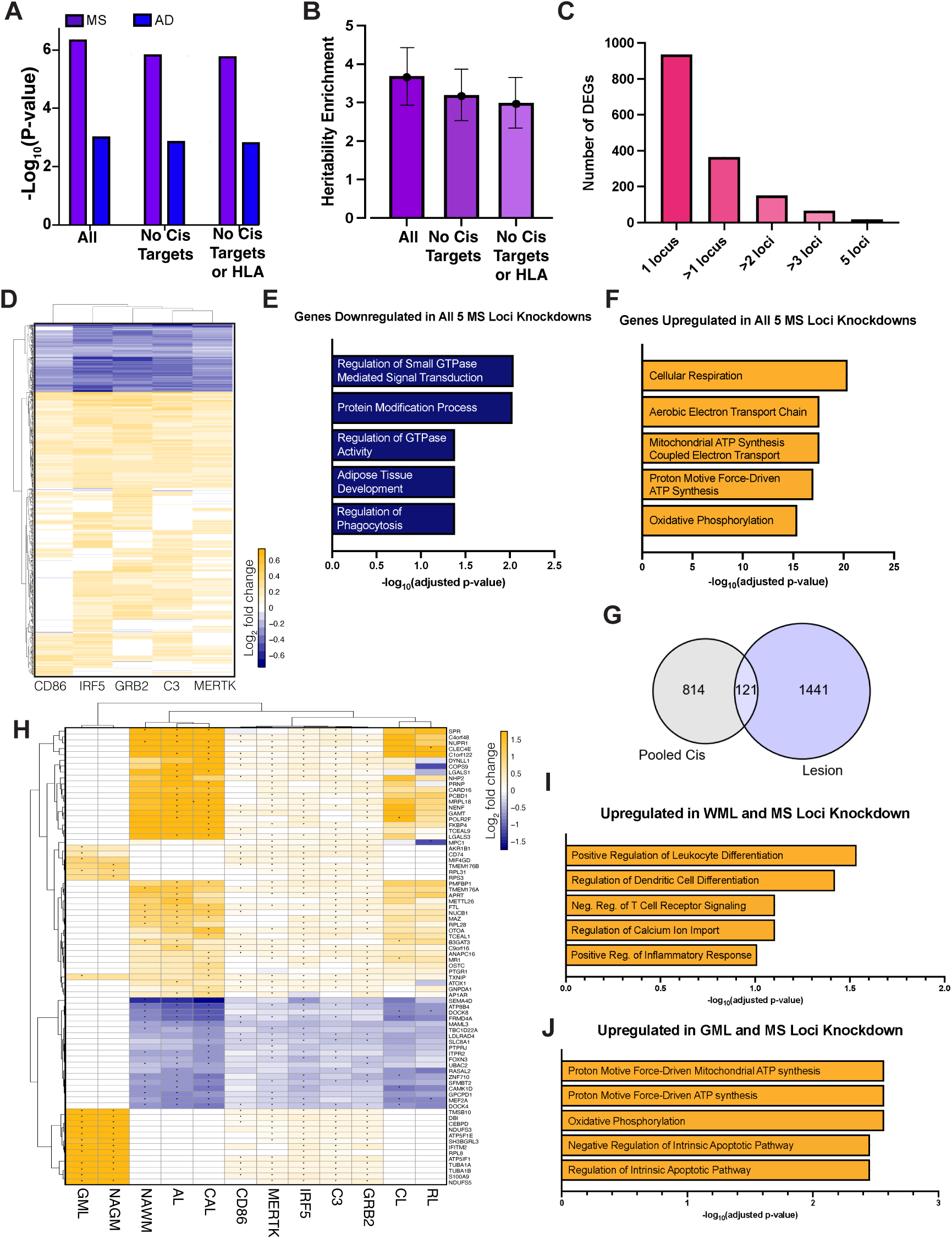
Downstream DEGs converge on shared biological pathways linked to microglia dysfunction in MS. **(A).** S-LDSC SNP heritability for Multiple Sclerosis (MS) and Alzheimer’s Disease (AD) is shown for 3 gene sets. “All” DEGs identified across top 5 loci (N=935); “No Cis Targets” all trans-acting DEG excluding the cis-acting gene targets (C3, CD86, GRB2, IRF5, MERTK) (N=930); and “No Cis Targets or HLA” trans acting targets excluding the HLA gene locus. **(B)** SNP heritability enrichments for MS, using the same categories as panel A. **(C)** Number of DEGs identified at one or more perturbed MS risk locus. **(D)** Heatmap of 935 DEGs found in at least one of the top 5 MS target loci. Genes that were not detected are designated a log2 fold change of 0. **(E)** Top five results for Gene Ontology (Biological Processes) analysis of downstream DEGs downregulated in all 5 MS loci knockdowns (168 genes); p-values from Enrichr. **(F)** Top five results for Gene Ontology (Biological Processes) analysis of downstream DEGs upregulated in all five MS loci knockdowns (260 genes); p-values from Enrichr. **(G)** Venn Diagram showing significant overlap between Perturb-seq DEGs and DEGs identified comparing MS patient microglia to control sample microglia N=1,562. (Fisher’s exact test P<0.001). **(H)** Heatmap of concordant DEGs across Perturb-seq and MS patient lesions. GML = gray matter lesion, NAGM = adjacent normal-appearing gray matter, NAWM = adjacent normal-appearing white matter, AL = active demyelinated lesion, CAL = chronic active demyelinated lesion, CIL = chronic inactive demyelinated lesion, RL = re-myelinated lesion. Asterisks indicate significant differences (corrected P<0.05) between MS patients and controls for patient data and significant differences between pooled-cis and pooled-control guides for each perturbed MS risk locus. **(I)** Top five results for Gene Ontology (Biological Processes) analysis of DEGs upregulated significantly in at least one MS locus knockdown and at least one gray matter lesion type (GML or NAGM) (20 genes); p-values from Enrichr. **(J)** Top five results for Gene Ontology (Biological Processes) analysis of DEGs upregulated significantly in at least one MS locus knockdown and at least one white matter lesion type (CAL, CIL, RL, or NAWM) (40 genes); p-values from Enrichr.

Finally, we compared the transcriptional effects of MS GWAS loci to transcriptional changes observed in MS cases. Single nucleus RNA-seq (snRNA-seq) was previously performed on brain tissue from MS cases as well as healthy control tissues [47]. In this study, MS DEGs (n=1,562) were identified by comparing microglia present in multiple lesion types as well as normal appearing gray and white matter found in MS tissues to healthy control samples. Of the 935 genes dysregulated as a result of MS risk gene perturbation, we found a significant number (n=121) to be differentially expressed in MS patient tissue microglia (Fisher’s exact test P<0.001). Even after restricting to directionally concordant DEGs, we observed a significant overlap (78/935, P<0.001, Fig 5G,H). Downstream DEGs that were found to be upregulated in MS patient gray matter lesions and normal appear gray matter were enriched for oxidative phosphorylation and proton motive force-driven ATP synthesis, similar to upregulated DEGs shared across our loc (Fig. 5J). Conversely, downstream DEGs that were upregulated in at least one MS patient white matter category (Fig. 5H) were enriched for immune cell differentiation and inflammatory response pathways (Fig 5I).

Collectively, these results demonstrate that the transcriptional consequence of MS risk loci predicted to act in microglia extend beyond risk SNP regulatory effects on *cis* target genes, and have highly convergent biological functions. The transcriptional changes identified in iMGL show strong concordance with MS genetic loci as well gene dysregulation observed in MS patients, linking both genetic risk pathways with pathophysiological alterations occurring after disease onset. This highlights the value of distinguishing the cell type of action of genetic risk loci, and performing perturbations on reduced numbers of select loci in order to better distinguish *cis* and *trans* effects with disease relevance. Overall, these approaches are well-suited to advance dissection of complex traits and reveal the underlying convergent mechanisms that drive disease pathogenesis.

## Materials & Methods

### iPSC culture

iPSCs reprogrammed from fibroblasts (Coriell Institute for Medical Research AG07657 and AG07599[6,16]) were cultured on mouse embryonic fibroblasts (MEFs), as previously described [16], for NPC differentiation. For all other purposes, cells were cultured on matrigel-coated plates (1:100 dilution in DMEM/F12) with mTeSR Plus, and passaged every 3-5 days with ReLeSR.

### iNPC, iPSC-N and iPSC-A differentiation

iNPCs were generated from iPSCs cultured on MEFs and maintained using a previously published protocol[13]. To generate neurons, iNPCs were co-transduced with 2nd generation lentivirus expressing murine *Ngn2* and *rtTA*, as previously described for iPSC transduction [48], after plating at 25,000 cells/cm2 in matrigel-coated dishes on D0. 1ug/mL doxycycline (dox) was added 24h later (D1) and media containing dox was freshly added every day until D7, after which dox was removed [14]. iPSC-N cultures were fixed for ICC analysis on D7,, and MEA analyses, RNA-seq and ChIP-seq were performed at D21. iPSC-A were generated from iNPCs using a published protocol [15]. Fixation for ICC, RNA-seq and ChIP-seq were performed after ∼8 weeks of differentiation.

### iMGL differentiation

iPSCs were cultured under feeder-free conditions and differentiated into iMGL according to previous studies [16]. Myeloid progenitors were collected from 10cm dishes and cultured in microglia maturation media (MGM) for 10 days before ICC, ChIP-seq, RNA-seq and Promoter Capture Hi-C were performed. Phagocytosis and lipid uptake assays were performed after 9 days in MGM.

### Immunocytochemistry

iNPCs, iPSC-N and iPSC-A were fixed with 4% paraformaldehyde for 20 min in matrigel/0.1% polyethyleneimine (PEI)-coated 24 well coverslip plates, iMGL were fixed in coverslips coated with 0.1% PEI only, and iPSCs were fixed in matrigel-coated plates. Fixed cells were washed in PBS+, permeabilized with 0.1% Triton X-100, blocked with 10% donkey serum, and incubated with primary antibodies overnight at 4C. The next day coverslips were washed with PBS+, incubated with secondary antibodies for 1.5-2h at RT in the dark, washed again, and stained with 0.2-0.5ug/mL DAPI for 3 min. After washing, coverslips were transferred to glass slides with Prolong Gold antifade mounting reagent and incubated overnight at RT in the dark, whereas iPSCs were imaged in the original plates. Epifluorescence microscopy was performed with Zeiss AxioPlan2, Nikon Eclipse Ti-U and Nikon Eclipse TE2000-U microscopes, confocal microscopy was performed with a Zeiss LSM700. Image analyses were performed with ImageJ.

### Multielectrode array analysis

Dox-treated and untreated *Ngn2*-transduced NPCs were passaged onto a 12-well CytoView MEA plate 21 days post-transduction at 500,000 cells per well. Neurobasal-based media containing 10ng/mL BDNF, 10ng/mL NT-3 and 1-2ug/mL mouse laminin A was changed every 3 days, and 30 minute recordings were performed 24h after each media change. Each plate was recorded four times with six biological replicates per condition.

### RNA-seq and data analysis

Total RNA was extracted using the QIAgen miRNeasy Mini Kit, libraries were prepared with the KAPA HyperPrep Stranded Kit, and samples were sequenced on a HiSeq lane with 100bp paired end reads. Reads were filtered with FastQC and aligned to the hg38 reference genome with TopHat and Cufflinks. For analysis of publicly available data, raw reads were downloaded from NCBI GEO and aligned with STAR19 (v2.7.1a) to the GRCh38 (hg38) human genome, canonical chromosomes only, using gene annotations from Ensembl Release 93 and the option ‘—sjdbOverhang 49’. Gene counts were obtained using featureCounts (v1.6.2) with Ensembl Release 93 gene models and converted to FPKM with the command ‘fpkm(robust = FALSE)’ from DESeq2 (v1.26.0). Normalized FPKM values were used to generate principal component plots, with or without batch correction by ComBat, using R package sva, v3.34.0). Data from human patients, including ex vivo brain cells, were obtained through dbGaP and analyzed similarly. Differentially expressed genes between cell types were identified using DESeq2 with a cutoff of >2-fold change and P<1e-05. Gene ontology analyses were performed on these genes using Metascape [17].

### Phagocytosis and lipid uptake assays

For phagocytosis, iMGL were treated with 25ug/mL or 100ug/mL pHrodo green-labeled zymosan beads for 1h, either with or without 1h pre-treatment with cytochalasin D. Mock treated cells received PBSwith matched volume of the higher bead dose. Cells were analyzed by flow cytometry for GFP, representing phagocytosed beads. For lipid uptake, iMGL were treated with 10ug/mL or 50ug/mL oxidized DiO-labeled low density lipoprotein for 2.5 or 4h, and were analyzed for GFP, representing intracellular lipid, by flow cytometry.

### ChIP-seq and data analysis

Cell pellets were prepared according to the Covaris TruChIP Chromatin Shearing kit, and sonication parameters were determined separately for each cell type. IP was performed with the Abcam ab4279 anti-H3K27ac antibody. ChIP libraries were prepared with a Swift ACEL-NGS 2S kit, and all samples were sequenced on a HiSeq lane with 50bp single end reads.

Reads were aligned with ‘bwa aln’ (v0.7.17-r1188) to the GRCh38 (hg38) human genome, canonical chromosomes only. Peaks were called using MACS221 (v2.2.7.1) with ‘—extsize’ determined by ‘phantompeakqualtools’ run_spp.R and the options ‘—keep-dup auto – nomodel’. Ex vivo brain cell data were accessed through dbGaP and analyzed in the same manner. Peaks enriched in, or unique to, different cell types were identified DiffBind, and heatmaps were also generated with DiffBind. The Integrative Genomics Viewer (IGV) was used to visualize signal and peaks at genomic locations.

### S-LDSC analysis

Heritability enrichment was estimated using linkage disequilibrium (LD) score regression [12], by comparing the peaks either unique to or enriched in a cell type of interest compared with other cell types from the same (ex vivo or iPSC-derived) source. LD scores and heritability enrichment calculations were performed using the baseline model composed of multiple functional categories. The significance threshold was defined through Bonferroni multi-test correction for the 51 tested traits. H3K27ac differential enrichment was calculated with DiffBind using the recommended default settings (http://bioconductor.org/packages/release/bioc/vignettes/DiffBind/inst/doc/DiffBind.pdf). PCA and heatmaps were also generated using DiffBind. Cell-type specific peaks were defined as peaks significantly enriched in a given cell type (padj<0.05) and not called in any other cell types.

### Promoter Capture Hi-C and data analysis

PCHi-C was performed using the Arima-HiC+ kit, Arima Human Promoter Capture Probes, and Arima Library Prep Module according to the manufacturer’s protocols. The capture probes target 18,741 protein-coding genes, 84 antisense RNAs, 170 lincRNAs, 1,878 microRNAs, 938 snoRNAs and 1,898 snRNAs. Briefly, 10 million iMGL were fixed after 10 days in MGM, aliquoted into separate 1 million cell aliquots, flash frozen in liquid nitrogen, and stored at −80C. One aliquot was determined to yield ∼3ug of DNA, which is ideal for PCHi-C, so a single aliquot was used for PCHi-C. The thawed aliquot was subjected to restriction enzyme digestion, ligation, library preparation, and sequenced on a NovaSeqS1 lane with 100bp paired end reads. Data were analyzed with the Arima Capture HiC Pipeline with modified HiCUP and CHiCAGO algorithms. Linear regression was used to determine the relationship between PCHi-C interactions and gene log2 mRNA levels, and Fisher’s exact test was used to test the relationship between PCHi-C interactions and H3K27ac peaks.

### Lentivirus generation

For *Ngn2* transductions of iNPCs, HEK293Ts were transfected with FUW-TetO-Ngn2-P2A-EGFP and FUW-M2rtTA transfer plasmids, along with pCMV-VSV-G envelope plasmid and pCMVR8.74 packaging plasmid, using X-tremeGENE 9. Virus-containing media was collected 48h later and pelletted for 1h45m at 23,000rpm with an ultracentrifuge. Pellets were resuspended in PBS, aliquoted and stored at −80C. For initial testing of the Vpx lentivirus system, pLentiCRISPR-EGFP was used as the transfer plasmid, whereas pBA904 was used for CRISPRi experiments and Perturb-seq., Transfer plasmids were co-transfected into HEK293Ts with pCMV-VSV-G, psPAX2-chp6 modified packaging vector, and pcVpx.myc [49], with X-tremeGENE 9. The next morning HEK293Ts were switched to IMDM +20% inactivated cosmic calf serum, and viral supernatant was collected in the afternoon 2 or 3 days post-transfection, and frozen at −80C.

### Generation of dCas9-KRAB iPSCs

Female iPSCs from AG07657 fibroblasts were transfected with pPX458-AAVS1-sg [50], which expresses Cas9, EGFP and a sgRNA targeting the *AAVS1* locus [51], and an *AAVS1*-targeting donor construct containing a dCas9-KRAB transgene, with the *ZIM3* KRAB domain fused to the dCas9 N-terminus, mCherry placed downstream of dCas9 and a P2A sequence, and neomycin/kanamycin resistance. Successful knock-ins were obtained by treating cells with 150ug/mL G418, and individual clones were characterized by expression of mCherry, OCT4, NANOG, Tra-1-160 and SSEA4. One clone with robust expression of all four pluripotency markers and mCherry was determined to be karyotypically normal and was [16] used for all CRISPRi experiments. Transgene insertion was independently verified by junction PCR.

### Lentiviral transduction of iMGL

Lentivirus produced with or without co-transfection of pcVpx.myc was titered on iMGL by plating 100,000 iMGL in MGM at 100,000 cells/mL onto viral supernatant in a total of 1mL IMDM, in 24 well plates. Each well had a final volume of 2mL, with 1mL MGM and 1mL IMDM plus virus. Media was aspirated the following morning and was replaced with 1mL MGM, and this was performed every 2 days until at least D7 post-transduction. For CRISPRi single guide knockdown experiments and Perturb-seq, cell numbers, lentivirus, and total volume were scaled up appropriately, maintaining the same cell density and media composition.

### Pro-inflammatory cytokine analysis

Media from transduced iMGLs was collected 3, 7 and 9 days post-transduction and frozen at - 80C. Upon thawing, samples were centrifuged for 5 min at 3000g, and the supernatant was submitted to Eve Technologies (Calgary, Alberta, CA) for duplicate analysis with the Human Cytokine Proinflammatory Focused 15-Plex Discovery Assay Array (HDF15).

### Flow cytometry

iMGL were dissociated with trituration or 0.25% Trypsin/EDTA, washed in FACS buffer, stained with eFluor780, washed again and filtered through a mesh cap into 5mL polystyrene tubes. When sorting cells for qPCR, BFP+ and BFP-cells were sorted with a 100um nozzle on a BD FACSAria II into MGM. For Perturb-seq, only BFP+ cells with the top 50% mCherry expression were sorted for scRNA-seq into PBS with 1% FBS and 25mM EDTA. Sorted cells were immediately spun for 5 min at 500g and resuspended in either RNA extraction buffer for qPCR, or a PBS-based scRNA-seq buffer for Perturb-seq.

### sgRNA design and cloning

For test knockdown experiments, published sgRNAs targeting the *B2M* and *CD151* transcriptional start site (TSS) [52], in-house sgRNAs targeting the *BIN1* and *ELMO1* TSS, and sgRNAs used to delete the AD risk SNP containing *BIN1* enhancer [10], were annealed and cloned into pBA904 using published methods [7]. sgRNAs were designed to target ideal TSS regions based on genome-wide CRISPRi experiments [10]. For Perturb-seq, TSS-targeting guides and non-targeting control guides were selected from a previous study [52] or by CRISPick, and selected highest ranked guides starting with a 5’ guanine. Enhancer-targeting guides and non-enhancer-targeting control guides were selected with guidescan2 (specificity >0.6 among top 10 ranked guides) or CRISPick. Additional negative controls sgRNAs targeting intergenic regions, intronic regions without H3K27ac, and regions with H3K27ac in non-microglial cell types only, were included. Finally, validated ELMO1 *TSS* guides from our test knockdown experiments and previously published *BIN1* enhancer targeting guides [10] were also added to the screen. The final screen design consisted of 387 unique 75bp oligos, each with a unique sgRNA protospacer, which were synthesized by Twist Biosciences. The sgRNA library was PCR amplified, cloned into pBA904 with NEB HiFi assembly, and transformed into NEB 5-alpha competent cells. Individual colonies were validated as containing expected protospacers by Sanger sequencing, and bacterial cultures were then expanded to perform Maxipreps. sgRNA representation in the library was assessed by PCR amplification of the protospacer region followed by sequencing on a single miSeq lane.

### Cloning of psPAX2-chp6 Vpx packaging plasmid

To generate psPAX2 plasmid with a chimeric Vpx packaging sequence, psPAX2 (a gift of Didier Trono) was digested with EcoRV and SphI and dephosphorylated, and pMDL-X (ref: Bobadilla et al 2013) (a gift of Nathaniel Landau) was digested with EcoRV and SphI. The digested vector and insert fragment were gel purified and ligated with T4 DNA ligase. Stbl2 cells were transformed with the ligation reactions, and correct clones were verified by Sanger sequencing. All enzymes were purchased from New England Biolabs.

### RT-qPCR

RNA extraction was performed with the RNeasy Plus Mini kit, and RT-PCR with qScript. qPCR was performed on cDNAs with Fast SYBR Green and primer pairs that were validated with a standard curve and melt curve analysis, requiring R2>0.98, efficiency of 90-110%, and a single melt curve product. GAPDH and ACTB were used as normalizers. For CRISPRi knockdown experiments, knockdown was determined by quantifying the sgRNA target gene expression levels in sorted BFP+ cells transduced with each sgRNA with BFP+ cells transduced with a non-targeting sgRNA.

### Differential Expression Testing

<u>Filtering:</u> Perturb-Seq analyses were performed in Seurat v5.2.1. Cells were filtered using the following metrics: >= 250 genes per cell, >= 500 UMIs per cell, <= 20% mitochondrial ratio, and >= 0.8 log_10_genes per UMI. Cells with 3 or more raw reads of a given gRNA were considered positive for that gRNA. Cells not transduced with any gRNA were excluded, yielding 65,906 cells total for downstream differential expression analyses. Gene expression data was scaled and centered using Seurat ScaleData.

<u>UMAP</u>: We ran UMAP on the first 5 principal components of the data. Using Seurat’s FindClusters with a resolution of 0.8, which yielded 13 clusters.

### Guide-specific DEGs

For individual guide analyses, cells with any combination of non-targeting control guides, but no other guides, were used as the control population (“NTC” – 1021 cells). Cells with any number of a given gRNA, regardless of other gRNAs’ status, were compared to the NTC population for guide-specific DEG testing. Differentially expressed genes were identified using a Wilcoxon Rank Sum Test (FindMarkers in Seurat). DEGs were then filtered for >= 15% fold change between groups, >= 10% expression in both groups, and a Benjamini-Hochberg corrected p-value < 0.05, yielding a filtered DEG list. We used EnhancedVolcano to visualize guide-specific DEGs.

### Locus-wide DEGs

Using guide-specific DEGs versus NTC, guides were designated as cis-targeting at a locus if the target gene was included in the filtered DEG list. Guides were designated as non-cis-targeting if the target gene was not included in the filtered or unfiltered DEG list. We pooled cells with only non-cis-targeting guides into a new control group (NoCis – 9091 cells). Then, we pooled all cells with cis-targeting guides targeting the same gene, and performed locus-wide comparisons versus NoCis. These DEGs were used for heat maps, comparisons to lesion data, and Gene Ontology as included in Figure 5.

### MS Patient Tissue snRNA-seq analysis

We utilized snRNA-seq data comparing MS patient lesions to control tissue, stratified by cell type [47]. When comparing DEGs, we included only genes that had a statistically significant fold change (p<0.05) in at least one lesion DEG list and one locus-wide DEG list. From here, we considered DEGs with a fold change > 1 in at least one white matter lesion (CAL, RL, CIL, WML, NAWM) DEG list and one locus-wide DEG list to be upregulated in white matter and our data. DEGs with a fold change > 1 in at least one gray matter lesion (GML, NAGM) DEG list and one locus-wide DEG list were upregulated in gray matter and our data. Analyses were also performed on downregulated genes; however, the lists were small with very little overlap. Shar

## Supplementary Figure Legends

**Figure S1: Transcriptomic and functional characterization of iPSC-derived cells. (A)** Dox-inducible *Ngn2*+ neurons display spontaneous synaptic activity 21 days after dox treatment, whereas NPCs never exposed to dox do not. **(B)** Neuronal, astrocyte and microglial marker gene expression in cells derived from 599 (male) and 657 (female) iPSCs. All panels except lower left display expression in TPM, whereas the lower left panel displays TPM normalized to the NPCs. **(C-F)** Gene ontology enrichments of top 300 most upregulated genes in the indicated cell types (NPCs, neurons, astrocytes and microglia, respectively), compared with the second indicated cell type (microglia, microglia, microglia and NPCs, respectively), using Metascape [17]. **(G)** PCA of RNA-seq analysis of 599- and 657-derived NPCs, astrocytes and microglia, and 657-derived neurons. Three biological replicates of 657 microglia are shown, as well as 657 myeloid progenitors, which were not exposed to microglial maturation media. **(H)** Flow cytometry analysis of iMGLs treated with two doses of FITC-labeled zymosan beads, with or without pre-treatment with cytochalasin A. Two replicates of each condition are shown; FITC signal represents bead phagocytosis. **(I)** Flow cytometry analysis of iMGLs treated with two doses of DiO-labeled LDL for two treatment durations. FITC signal represents lipid uptake. For panels H and I, untreated cells were stained with the viability dye eFluor 780 alone.

**Figure S2: Comparison of H3K27ac-defined enhancers in ex vivo and iPSC-derived cells. (A)** H3K27ac ChIP-seq was performed on unsorted iPSC-derived NPCs, neurons, astrocytes and microglia, and compared with ChIP-seq data from NeuN-, LHX2-, PU.1- and OLIG2-sorted nuclei from fresh brain tissue (PMID 31727856). These data were then used for s-LDSC analyses in Fig. 2. **(B-D)** UCSC and IGV browser snapshots of H3K27ac signal from ex vivo and iPSC-derived cells, respectively. In each panel, ex vivo data are displayed on top and iPSC-derived data are below. *NEUROD2*, *ALDH1L1* and *SPI1* are shown as examples of neuron-specific, astrocyte-specific and microglia-specific loci, respectively.

**Figure S3: Transcriptomic and epigenetic characterization of iMGLs and comparison to published iMGL and ex vivo microglia. (A)** PCA showing first two principal components of RNA-seq data from four published iMGL protocols (red, dark brown, yellow and light brown), iMGLs from this paper (black), ex vivo microglia (blue) and ex vivo microglia after cell culture (green). Individual sample are labeled by dataset, microglia type, treatment, biological replicate and technical replicate (e.g. Dou_iPSC_MG1_2 represents technical replicate 2 of biological replicate 1 from the Dou iPSC dataset. **(B)** PCA showing first two principal components of RNA-seq data from iPSC-derived cells from this study (black), ex vivo microglia with (green) and without cell culturing (blue), and an independent ex vivo microglia dataset (red). **(C)** Enrichr gene ontology analysis of enhancers present in ex vivo microglia but absent in iMGLs from this study, with loci containing the highest number of ex vivo-specific H3K27ac peaks listed on the right. **(D)** Distribution of the number of PCHi-C interactions per promoter, with the total, mean and median interactions listed. **(E)** Boxplot of PCHi-C *cis* (intrachromosomal) interaction distances. Median interaction distance is ∼250kb. **(F)** Boxplots of all expressed iMGL mRNAs split into TPM quintiles (0-20% is lowest quintile, 81-100% is highest quintile), with number of PCHi-C interactions for each gene displayed on the y-axis. The p-value represents the statistical significance of the correlation between mRNA levels and PCHi-C interactions when all genes are analyzed. **(G)** Metascape cell type gene signature sets most significantly associated with the 300 genes with the most PCHi-C interactions. Microglia gene signature sets are indicated by red arrows.

**Figure S4: Generation of dCas9-KRAB iPSCs and transduction of dCas9-KRAB iMGLs with Vpx lentiviral system. (A)** Normal female karyotype observed in the dCas9-KRAB iPSC clone used in this study. **(B)** Junction PCR identifies dCas9-KRAB transgene integration site at the *AAVS1* locus (1kb band) in knock-in clone (+dCas9) but not parental 657 iPSC (-dCas9) gDNA. H2O lane represents water control. **(C)** Flow cytometry analysis of mCherry expression in dCas9-KRAB (ZIM3) iPSCs compared with parental 657 iPSCs. **(D)** ICC analysis of dCas9-KRAB expression in transgenic iPSCs (bottom) and parental iPSCs (top). ICC was performed with an anti-Cas9 antibody and green secondary antibody; DAPI is blue. **(E)** Schematic of Vpx lentiviral system. HEK293Ts are transfected with transfer, packaging and envelope plasmids, with or without Vpx expression plasmid. pLentiCRISPR-EGFP was used as the transfer plasmid, and EGFP was visible only when Vpx was included. Cells were imaged 12 days post-

transduction, and differences in cell confluency are random and not related to transduction or presence of Vpx. **(F)** Pro-inflammatory cytokine ELISA performed on iMGLs with (+LV) or without (Mock) Vpx-mediated transduction of pLentiCRISPR-EGFP, both 3 and 9 days post-transduction. Cytokine levels are normalized to the mock D3 condition. **(G)** Morphology and BFP observed in iMGLs transduced with the BFP-expressing Perturb-seq vector, compared with untransduced cells, D9 post-transduction.

**Figure S5: Expression of microglia and macrophage marker genes in iMGLs transduced with Perturb-seq library.** Feature plots showing single cell expression of microglial markers *TREM2*, *AIF1 (IBA1)* and *TMEM119*, and brain border-associated macrophage marker *LYVE1*.

**Figures S6: Example MS GWAS locus ELMO1 with no distal sgRNA effects on cis targets.** Overview of ELMO1 locus shows H3K27ac ChIP-seq data for immune and CNS cell types (top), and locations of MS GWAS loci and outside variants stratified by risk (bottom).

**Figure S7: Comparison of pooled guide RNA analysis strategies.** Clustered heatmaps show comparisons of DEGs across different Perturb-seq analysis approaches. TSS vs NTC indicates the highest fold-change TSS guide vs cells that only received non-targeting guides, Enh vs NTC indicates the highest fold-change distal enhancer guide vs cells that only received non-targeting guides, and pooled cis vs pooled control indicates analysis performed using all cells with a sgRNA that impacts cis target gene (pooled cis) compared to all cells receiving only sgRNA that had no effect on cis-target (pooled control). For GRB2, no TSS sgRNA was available for comparison.

## Data Availability

All data produced in the present study are available upon reasonable request to the authors

