## Supplemental Figures for "Functional genomic dissection of MS risk loci reveals convergence of cis and trans gene regulatory mechanisms in microglia"

### Supplemental Figure 1

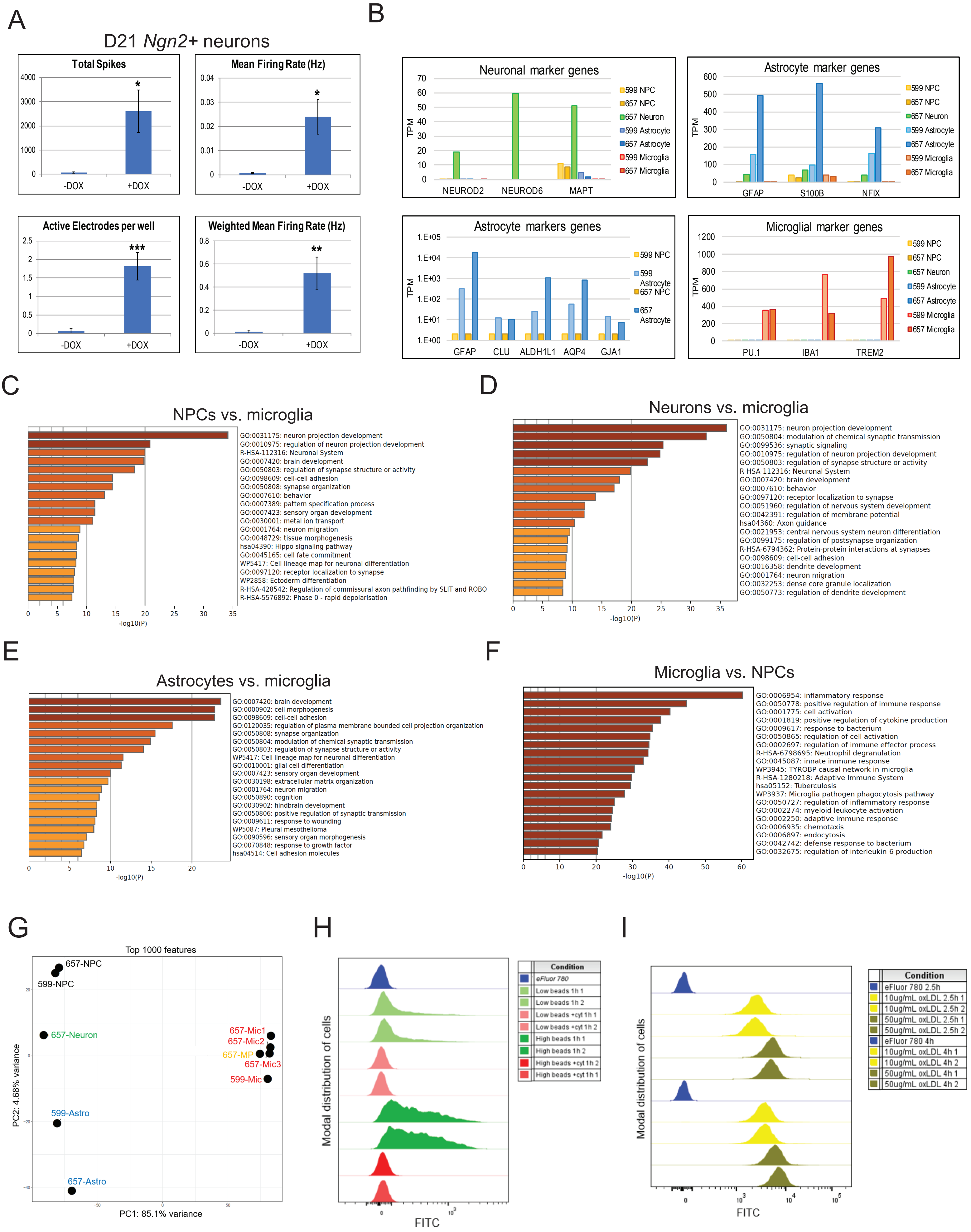

### Supplemental Figure 2

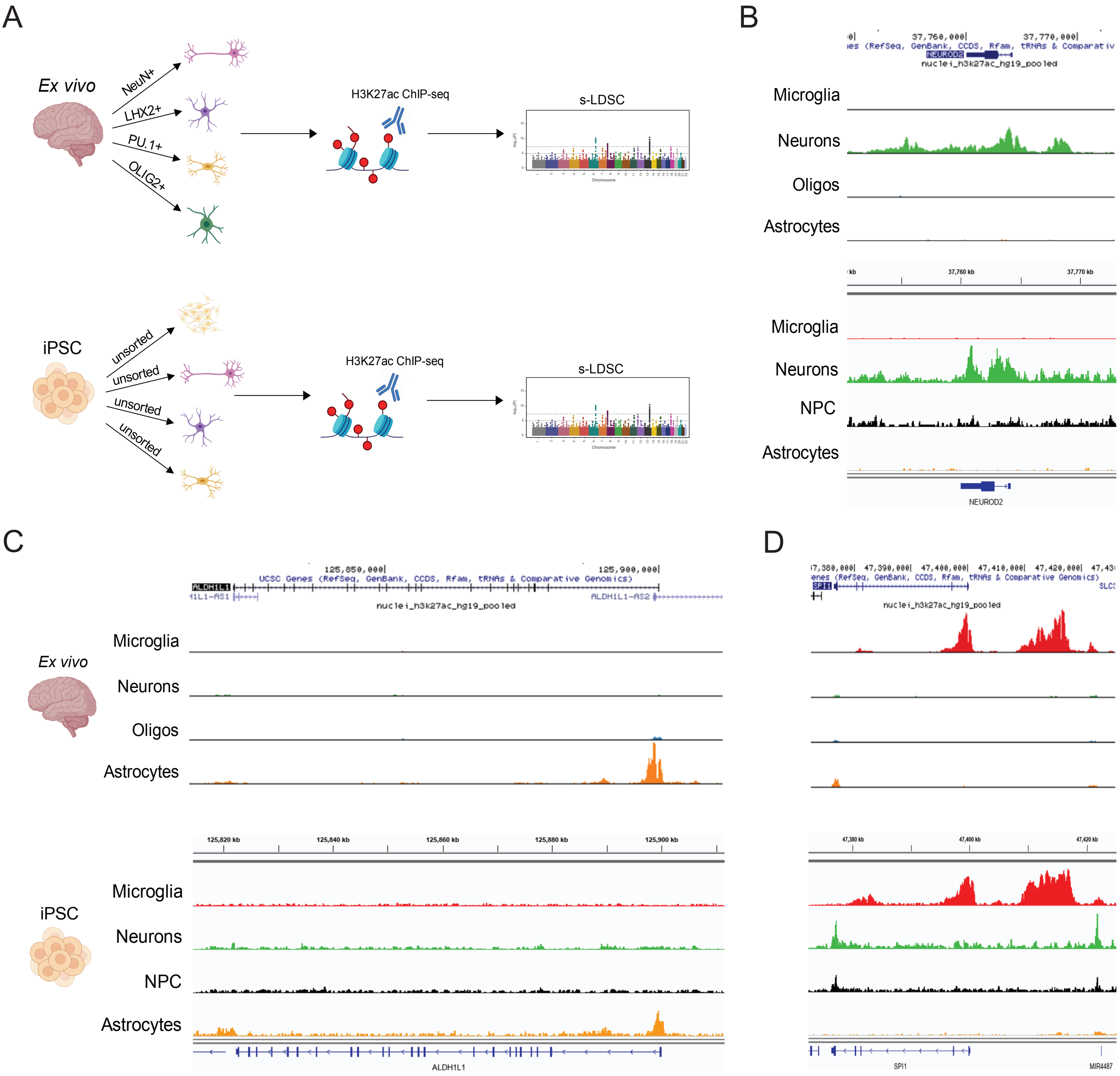

### Supplemental Figure 3

A

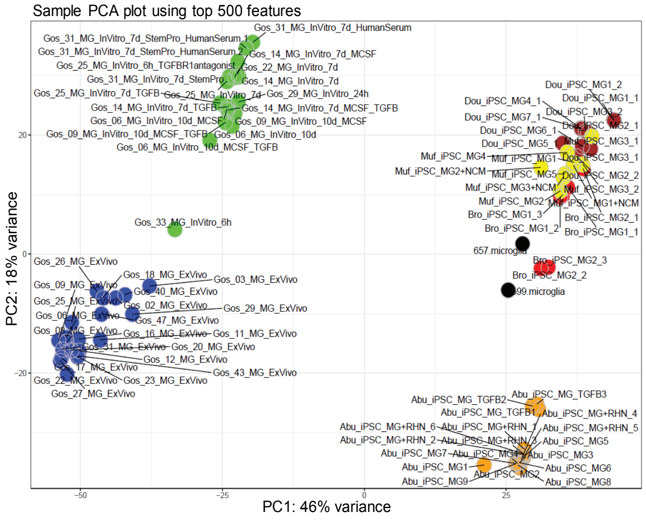


B

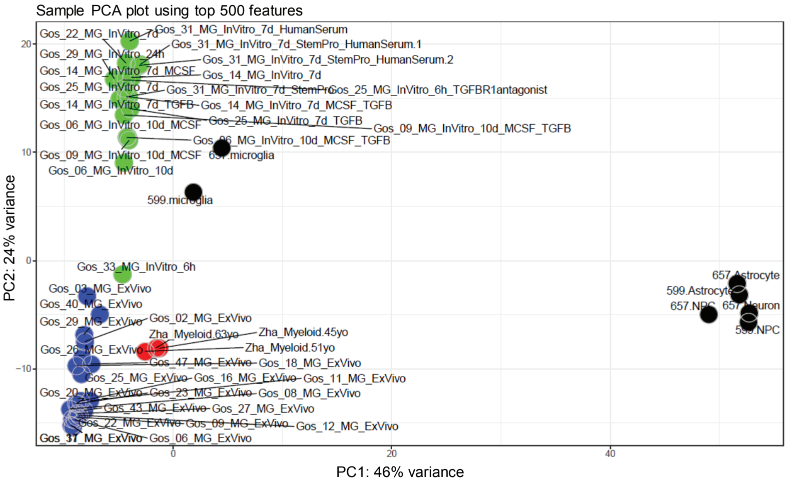


C

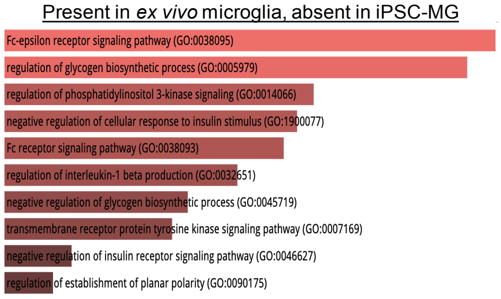


| # of peaks lost | PLAC-seq assigned gene |
|---|---|
| 315 | SALL1 |
| 88 | FGF20 |
| 62 | KRTPAP5-6 |
| 51 | PRKACG |
| 51 | PCDHGA12 |
| 44 | DKK1 |
| 42 | KCNQ3 |
| 40 | FAM59A |
| 38 | NKX3-1 |
| 37 | MRPS23 |

D

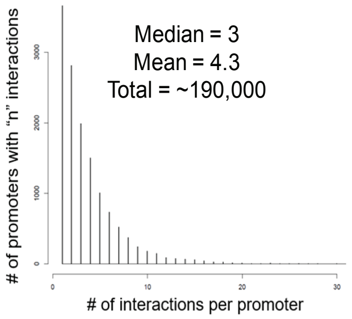


E

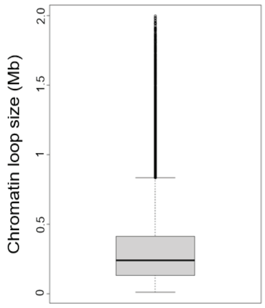


F

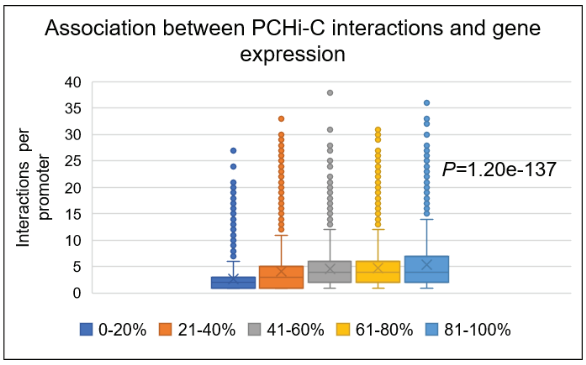


G

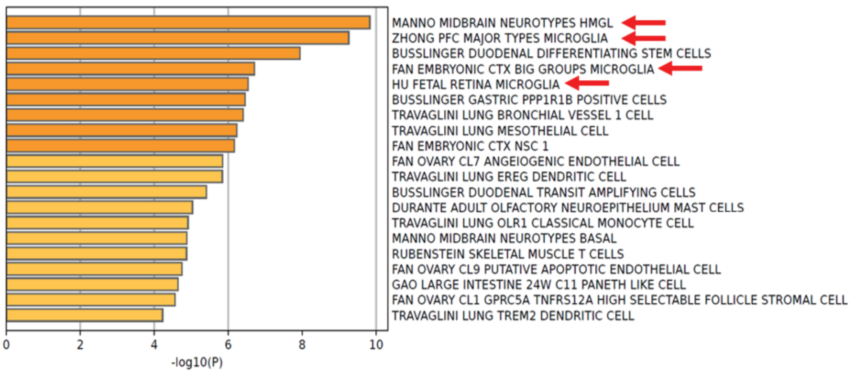

### Supplemental Figure 4

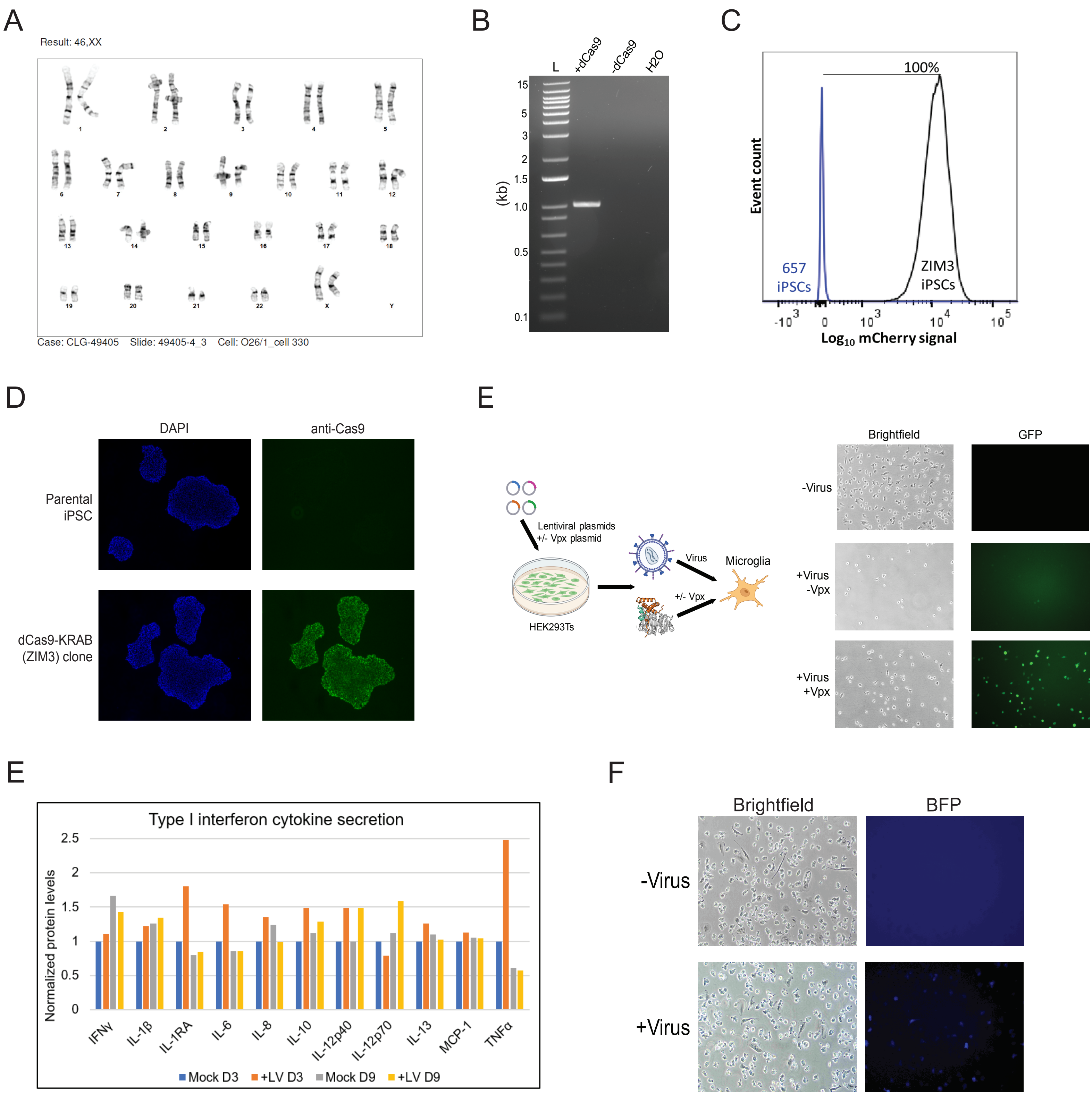

### Supplemental Figure 5

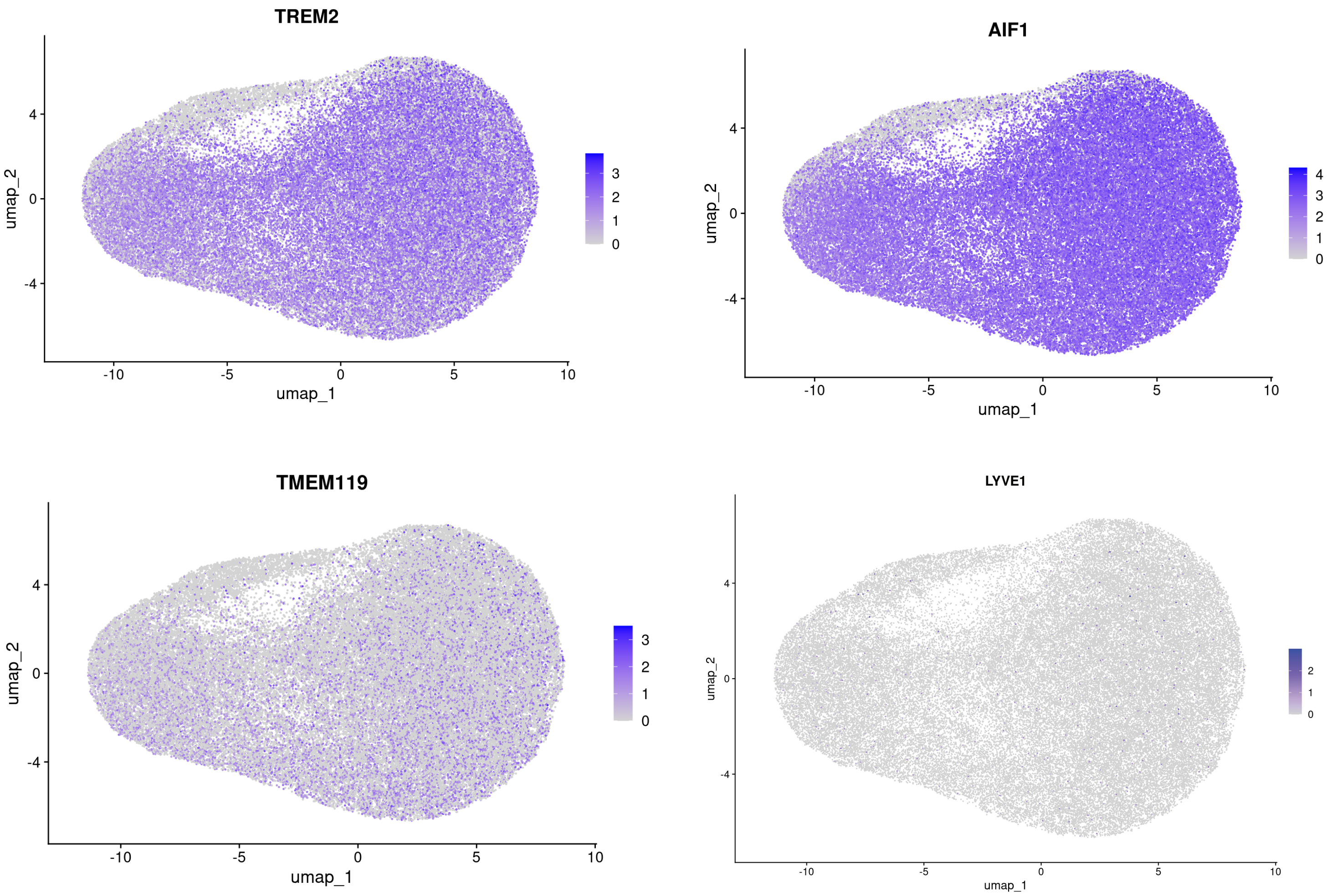

### Supplemental Figure 6

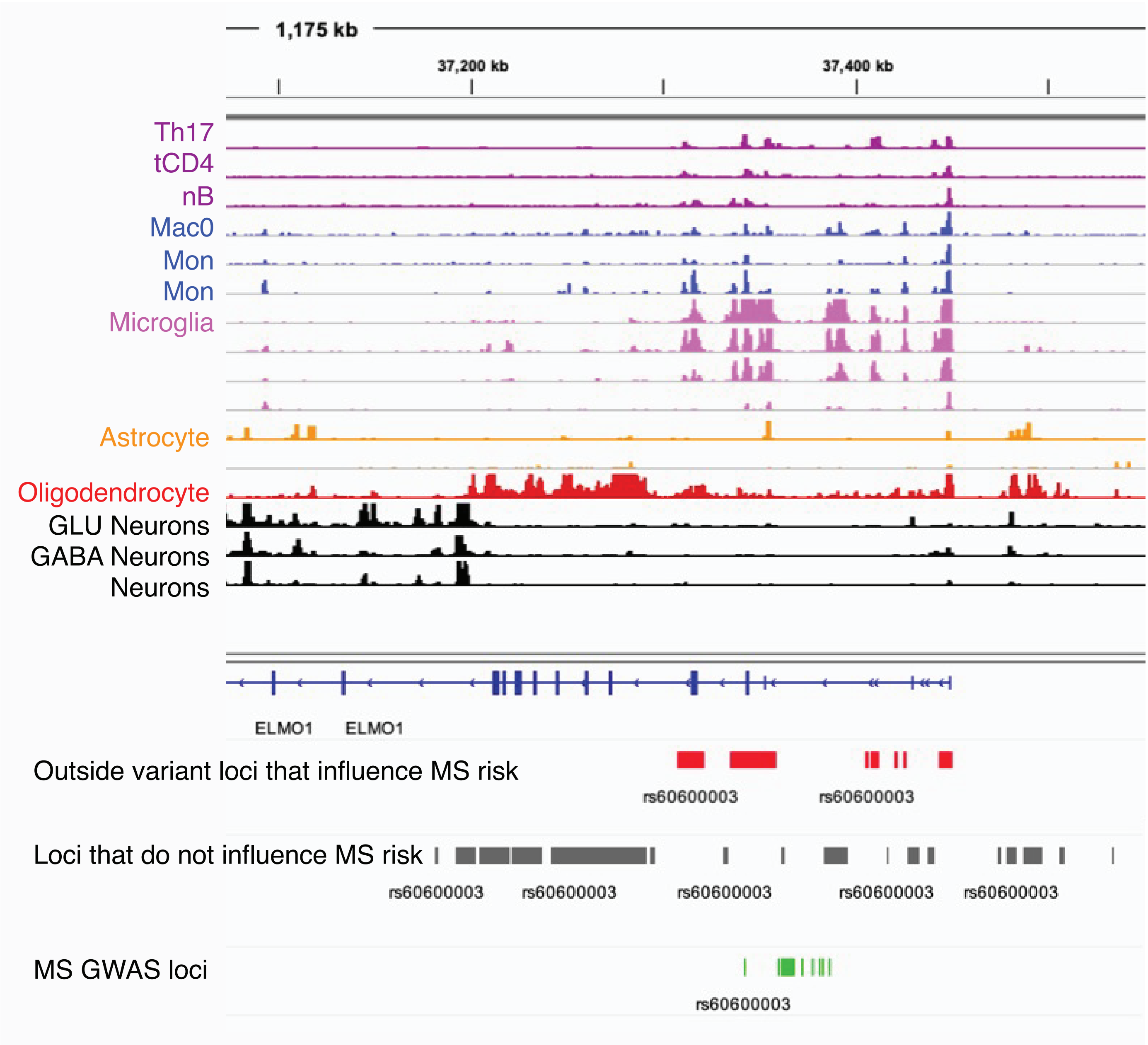

### Supplemental Figure 7

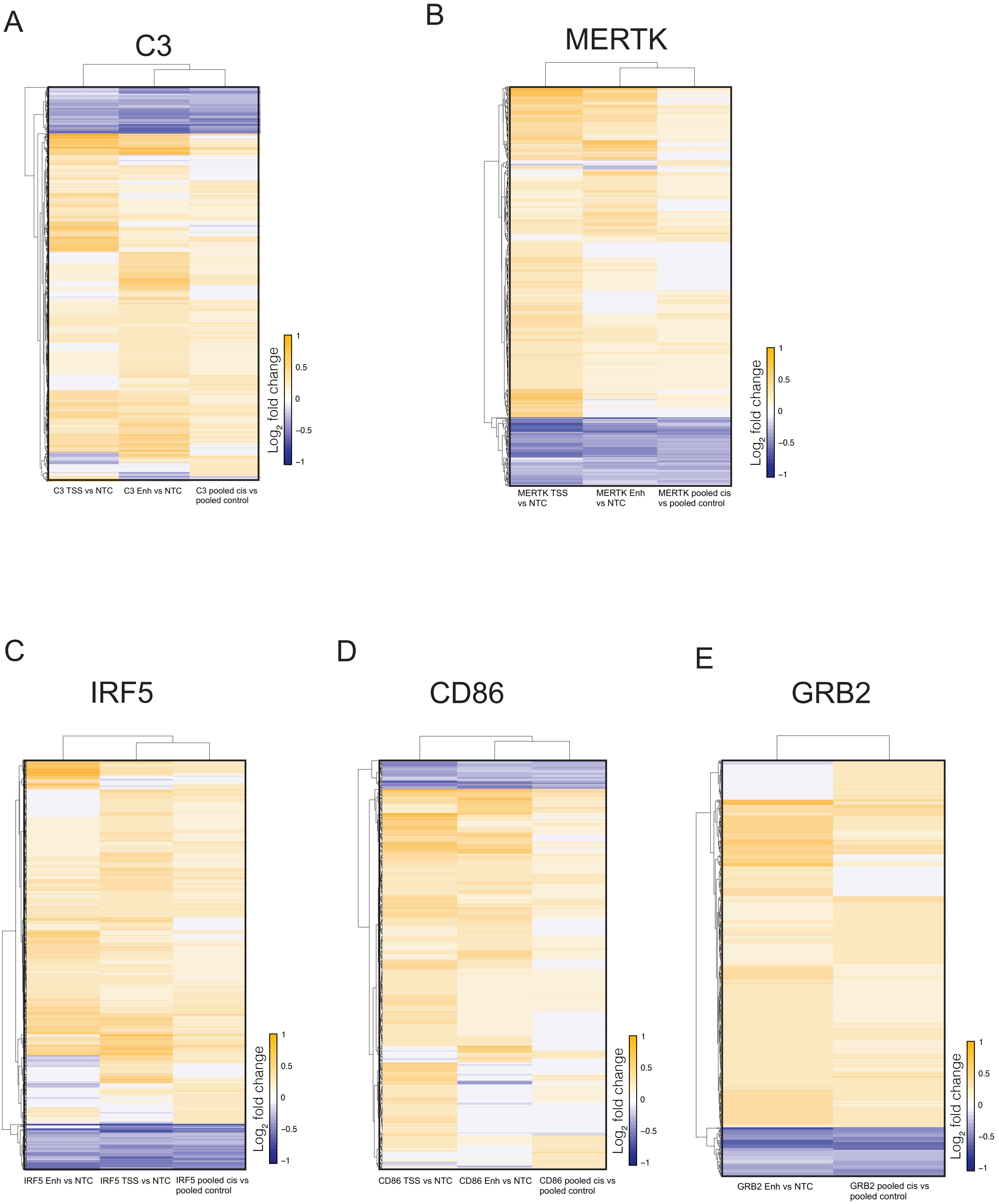
